# Private Health Sector in India: Ready and willing, yet underutilized in the Covid-19 pandemic

**DOI:** 10.1101/2020.06.09.20126086

**Authors:** Samira Davalbhakta, Supriya Sharma, Shefali Gupta, Vishwesh Agarwal, Gaurav Pandey, Durga Prasanna Misra, Bijaya Nanda Naik, Ashish Goel, Latika Gupta, Vikas Agarwal

## Abstract

**Background:** The private medical sector is a resource that must be estimated for efficient inclusion into public healthcare during pandemics.

**Methods:** A survey was conducted among private healthcare workers to ascertain their views on the potential resources that can be accessed from the private sector and methods to do the same.

**Results:** There were 213 respondents, 80% of them being doctors. Nearly half (47.4%) felt that the contribution from the private medical sector has been suboptimal. Areas suggested for improved contributions by the private sector related to patient care (71.8%) and provision of equipment (62.4%), with fewer expectations (39.9%) on the research front. Another area of deemed support was maintaining continuity of care for non-COVID patients using virtual consultation services (77.4%), tele-consultation being the preferred option (60%). 58.2% felt that the Government had not involved the private sector adequately; and 45.1% felt they should be part of policy-making.

**Conclusion:** A streamlined pathway to facilitate the private sector to join hands with the public sector for a national cause is the need of the hour. Through our study, we have identified gaps in the current contribution by the private sector and identified areas in which they could contribute, by their own admission.

## Introduction

The novel coronavirus disease (COVID-19) has consumed and exhausted widespread national health resources with unprecedented speed, and is expected to leave lasting consequences on global health, economy and growth. ^1^ The massive losses ^2^ call for the amalgamation of rapid innovations alongside bold public health measures led by a courageous political will to tackle this unique *“War sans Weapon”* situation. ^3^ As of May 28^th^ 2020, India has reported 158,332 COVID-19 cases, ^4^ a number that is rapidly rising, consuming the public healthcare system, which has been at the fore of this pandemic, despite deficient infrastructure, manpower and poor resources. ^5,6^ Amongst other countries, India currently ranks fourth with regards to daily increase in the cases. ^7^ With an availability of 0.55 public-hospital beds to 1000 population ^8^, it is not unreasonable to expect that the public sector may not be able to provide effective, sustained and uninterrupted healthcare in the face of the rising numbers. Not surprisingly, countries ahead of us on the pandemic curve have recognized the need to utilize all available healthcare resources, forging partnerships between public and private healthcare sectors.

In India, the healthcare scenario has transformed over the last few decades, ^9,10^ and almost 87% services are provided in the private sector, making it a major stakeholder. ^11^ The first decade of this century saw a growth in private sector beds by almost 70%, bringing their total share to nearly 63%.^12^ Although healthcare professionals in private enterprises are best suited to provide insights into potential areas of access from the private sector and methods to do so, yet there voices are seldom heard in the scientific world. Improvements in outcomes and health indicators have been reported after private-public partnerships (PPP) in previous reports. ^13^ The National Health Policy (NHP) 2017 not only advocates for exploring role of PPP in achieving Universal Health Coverage (UHC),^14^ but PPP has also been proposed as an efficient model for disaster risk reduction. ^15,16^

The present survey was conducted to explore the opinions and preparedness of healthcare workers (HCWs) in the private sector, on public-private partnerships (PPP) to provide a sustained, uninterrupted healthcare response in the face of the current pandemic.

## Methods

### Study population selection

An online survey was conducted in April 2020, and a pre-tested, content validated questionnaire was circulated over WhatsApp® groups of healthcare professionals (doctors, nurses, technicians, students and administrators amounting to nearly 2000 individuals) in the private hospitals across India.^17^ The participants were requested to provide an informed consent at the beginning of the survey. We did not offer incentives for participation.

### Questionnaire design

The questionnaire featured 20 questions, of which five identified respondent characteristics. Fourteen items were multiple choice, with one being open-ended. The average time to complete the survey was five minutes. The respondents could change the answers before submission but not after it. Internet Protocol addresses were checked to avoid duplication of responses. Content validity of the survey questionnaire was performed using Lawshe’s method and confirmed by three professors and one undergraduate medical student. The validated survey questionnaire was pre-tested among five HCWs, and the identified errors in wording, grammar or syntax were rectified. The ‘Logics’ feature available on Survey Monkey® allowed respondents to skip to a specific question on a later page based on their answer to a previous close-ended question. Descriptive statistics were performed, and the results were expressed as numbers (percentages). The data/figures were downloaded from surveymonkey.com®.

#### Ethics approval

Exemption from review was obtained from the institute ethics committee [2018-62-IP-EXP] as per local guidelines. We adhered to the Checklist for Reporting Results of Internet E-surveys to report the data. ^18^

## Results

The participants included doctors (80% of the 213 respondents) (age 35 years ± 11.1), and nurses or medical students (20%). Nearly half (47.4%) felt that the contribution from the private medical sector has been suboptimal. Suggestions for improved contributions included patient care (71.8%) and provision of equipment (62.4%), and research (39.9%). (Figure 1)

**Fig.1.**
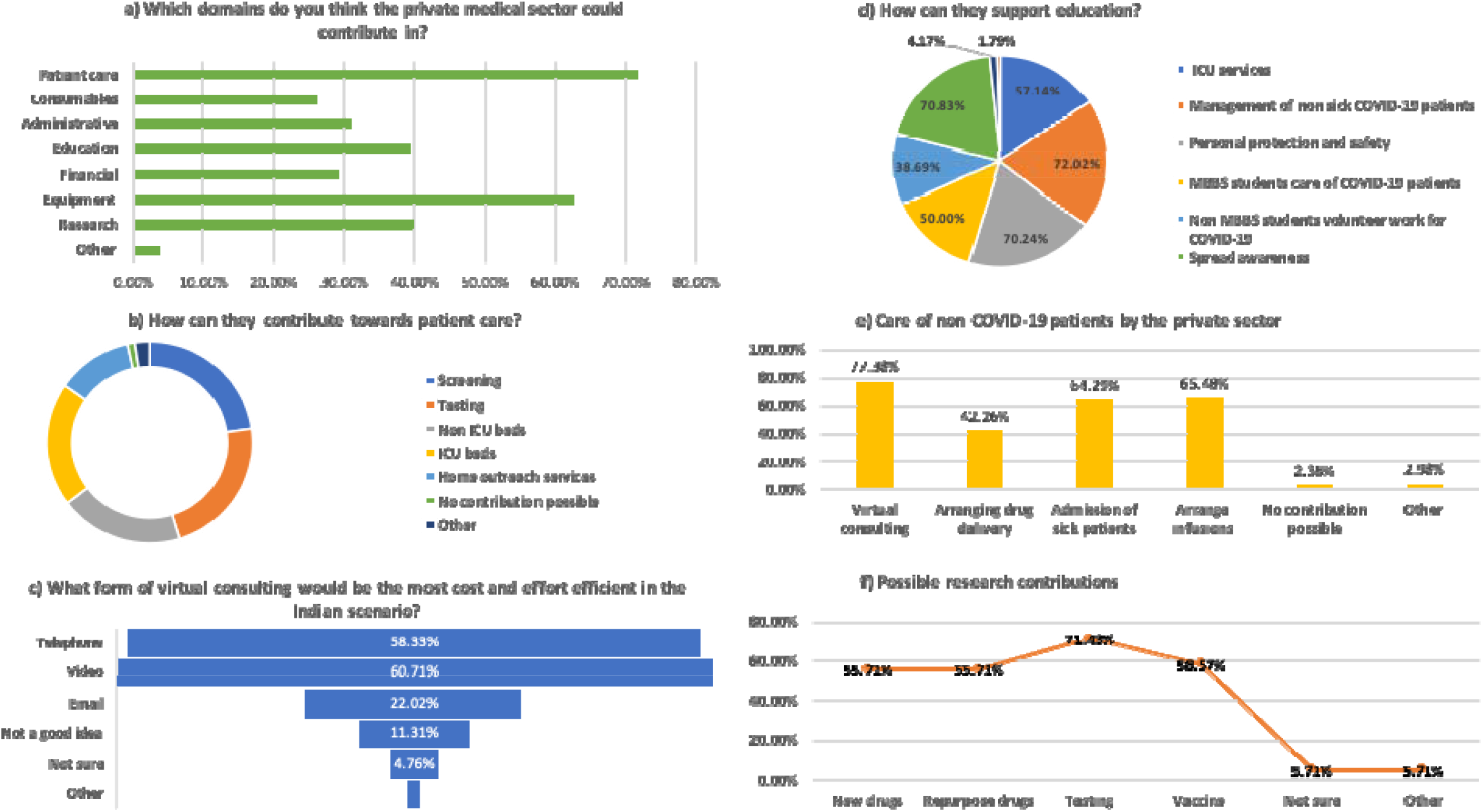
Responses from our survey. a)Domains that the private sector could contribute in. b)How they can contribute towards patient care. c) Form of virtual consulting that would be the most cost and effort efficient in the Indian scenario. d) How they can support education. e) How they can care for no COVID-19 patients. f) Research contributions

Participants suggested increased involvement in screening (69.6%), testing (69.1%), intensive care (61.31%) and non-intensive-care (59.52%) beds. Some (36.3%) felt that effective home outreach services could also be provided.

Participants believed that the private healthcare sector could provide insights into new testing methods (67.2%), vaccines (60.3%) and new or repurposed drugs (55.2%). Most participants (67.3%) preferred use of their financial contribution for subsidized treatment of patients while only 31% favored donation to public agencies.

Most respondents felt that they could play a significant role in educating healthcare workers, medical students, and the community. Another area of deemed support was maintaining continuity of care for non-COVID-19 patients, using virtual consultation services (77.4%), teleconsultation being the preferred option (60%).

More than half (58.2%) felt a need for greater involvement of the private sector in the pandemic response including policy making. Nearly half of the participants had made monetary donations for the pandemic from their personal funds (48.2%). Teleconsultations were being offered by 13 doctors. More than a third (37.36%) felt that they wanted to contribute more towards the pandemic response, and as many as 75% were keen to donate blood.

## Discussion

In our e-survey assessing the opinions and readiness of HCWs in the private healthcare sector, we found that participants felt that they had not contributed enough and were positively inclined to participate in the pandemic response. They expressed readiness to participate in screening, testing, patient care, support for equipment and clinical trials of newer drugs as well as repurposed medicines, vaccines or newer diagnostic tests.

While testing and tracing contacts remains the primary public health response to an infectious disease pandemic, over 3 million samples have been tested in India since January 2020 ^19^. Although we have attained testing capacity of 1 lac samples per day, it would still take more than three and a half years to test10% of the population. This appears to be an optimistically conservative but inadequate strategy in a country with more than 1.3 billion susceptible individuals. ^20,21^ Collaborations between government and private healthcare centers can decentralize screening and testing facilities, offloading central agencies while increasing the capacity and outreach.

While the public sector has been holding forte in the past few months, the need for additional resources is being increasingly felt. The private healthcare sector has significant potential,^22–24^ with 58% of the hospitals, 29% of the beds and 81% of doctors.^25^ Under severe strain, similar collaborations have been forged in Italy, Spain and several other countries.^26 27^ A similar exercise in India would be a prudent way ahead in these times.

A large number of blood banks are in the private healthcare sector in India, and it might be worthwhile to explore the conversion of private blood banks into specialized units for the promising convalescent plasma donation therapy, if efficacy is proven in ongoing clinical trials. This will not only tide over the ongoing acute shortage of blood products but also be a sustainable source of convalescent plasma for therapy in severe COVID-19. In fact, a vast majority of respondents expressed their willingness to donate blood to tide over the acute shortage of blood products in present times.

While most public facilities are busy in COVID-19 care, patient with *non-COVID* ailments have faced neglect and apathy. Private healthcare respondents are willing and prepared to participate.

Additionally, a forward triage protocol using tele-medicine services may in-fact hearald a revolution for a large number of technology-enabled non-COVID patients. In the Western world too, teleconsultations are being increasingly preferred as means of avoiding congestion in public spaces. ^28^ Although lower literacy levels and traditional patterns of doctor-patient interactions are a challenge in providing effective home-based outreach care in India, yet the scope of mobile networks and empowerment by these hand-held computers cannot be underestimated.^29,30^

Nearly two-thirds of the respondents felt that the private sector could leverage its financial resources by providing free or subsidized treatment to patients. While the Government makes efforts to meet the requirement of ventilators, stuttering from the onslaught of paused supply from Europe and China, it is prudent to recognize and utilize the dormant resources lying in the private hospital intensive care. ^31–33^ Further, private laboratories and research facilities, encouraged to develop new cost-effective and rapid high through-put testing methods, should start showing results soon.

In unusual times such as this, lessons could be learnt from past experiences. During the influenza pandemic of 2009, all dealings in India were restricted to the public sector to keep track of cases while ensuring affordable healthcare. This eventually led to an infrastructural deficit, and the consequent need to amend policies to include support from the private sector.^34^ Such experiences from previous epidemics have probably contributed to our HCWs believing that the private sector in India can participate in a more significant manner.

### Strengths and limitations

This is the first survey including a wide set of stakeholders in the private healthcare sector and in our opinion, it is an important move in the right direction to ascertain willingness and preparedness.

The results, subject to opinionated biases of a small set of young technology-empowered respondents, largely doctors are nevertheless enlightening and encouraging. Further, since this survey was electronically distributed, it has the advantage of a diverse representation of voices across the country, and yet opinions may be influenced by differential approach determined by local state policy.

## Conclusion

In the face of an unprecedented disease, with mystical transmissibility and unprecedented ability to devastate the human population, it is not surprising that the public healthcare sector is under more stress than it can handle alone. We have a large private healthcare sector in our country which is not only equipped but also willing, to share the burden of disease. Thus, a pragmatic approach to facilitate the private-public partnership will go a long way in mitigating the community impact and reduce mortality in current times. An open, healthy and swift discussion between the public and private sector should be the first step towards sorting grey areas.

## Data Availability

All data is present in the form of tables attached in the manuscript.

---

**Table 1:** Survey responses.

| **Table 1: Survey responses** |  |
| --- | --- |
| Question | Response [n (%)] |
| **Q1. Do you think the private medical sector in India is contributing enough towards the COVID-19 pandemic? [single response, N=213]** |  |
| No | 101 (47.42%) |
| Yes | 74 (34.74%) |
| I am not sure | 38 (17.84%) |
| **Q2. Which domains do you think the private medical sector could contribute in?[multiple responses, N=213]** |  |
| Research | 85 (39.91%) |
| Patient care | 153 (71.83%) |
| Financial | 62 (29.11%) |
| Education | 84 (39.44%) |
| Administrative (e.g. Formulation and execution of guidelines) | 66 (30.99%) |
| Consumables | 56 (26.29%) |
| Equipment (e.g. ventilators) | 133 (62.44%) |
| Other (please specify) | 8 (3.76%) |
| **Q3. How do you think the private sector could contribute towards research?[multiple responses. N=70]** |  |
| Develop new drugs | 39 (55.71%) |
| Repurpose existing drugs | 39 (55.71%) |
| Develop new methods of testing | 50 (71.43%) |
| Develop a vaccine | 41 (58.57%) |
| Not sure | 4 (5.71%) |
| Other (please specify) | 4 (5.71%) |
| **Q4. How do you think the private sector could contribute towards patient care?[Multiple responses, N=168]** |  |
| Screening services | 117 (69.64%) |
| Testing | 116 (69.05%) |
| Offer non ICU beds for admission | 100 (59.52%) |
| Offer ICU beds for admission | 103 (61.31%) |
| Start home outreach services | 61 (36.31%) |
| I don't think they can contribute in this domain | 6 (3.57%) |
| Other (please specify) | 12 (7.14%) |
| **Q5. How do you think the private sector could contribute towards consumables/supplies?[Multiple responses. N= 168]** |  |
| Masks | 101 (65.48%) |
| Drugs | 89 (52.98%) |
| Testing kits | 103 (61.31%) |
| Gloves and protective gear | 115 (68.45%) |
| Not sure | 9 (5.36%) |
| I don't think they can contribute in this domain | 28 (16.67%) |
| Other (please specify) | 7 (4.17%) |
| **Q6. How do you think the private sector could contribute financially?[Multiple responses, N=168]** |  |
| Contribution directly to local govt hospitals | 51 (30.36%) |
| Provide free or subsidised treatment to those who cannot afford it | 113 (67.26%) |
| Contribution to the PM fund | 43 (25.60%) |
| I don't think they can contribute in this domain | 20 (11.90%) |
| Other (please specify) | 9 (5.36%) |
| **Q7. How do you think the private sector could contribute towards education?[Multiple responses, N=168]** |  |
| Educating non ICU health care workers towards ICU services | 96 (57.14%) |
| Training of healthcare professionals for management of non sick COVID-19 patients | 121 (72.02%) |
| Training of other healthcare professionals towards personal protection and safety | 118 (70.24%) |
| Training of students (MBBS) towards volunteer work for COVID-19 | 84 (50.00%) |
| Training non MBBS students towards volunteer work for COVID-19 | 65 (38.69%) |
| Spread awareness amongst the population | 119 (70.83%) |
| I don't think they can contribute in this domain | 7 (4.17%) |
| Other (please specify) | 3 (1.79%) |
| **Q8. How do you think the private sector could contribute towards care for non COVID-19 patients?[Multiple responses, N=168]** |  |
| Virtual consulting | 130 (77.38%) |
| Arranging drug delivery | 71 (42.26%) |
| Admission of sick patients | 108 (64.29%) |
| Arrange infusions for those due for infusions (e.g. blood transfusion, vaccines, biologics, cancer chemotherapy) | 110 (65.48%) |
| I don't think they can contribute in this domain | 4 (2.38%) |
| Other (please specify) | 5 (2.98%) |
| **Q9. What form of virtual consulting do you think is the most cost and effort efficient in the Indian scenario?[Multiple responses, N=168]** |  |
| Telephone | 98 (58.33%) |
| Video | 102 (60.71%) |
| Email | 37 (22.02%) |
| I don't think virtual consulting is a good idea | 19 (11.31%) |
| I am not sure | 8 (4.76%) |
| Other (please specify) | 2 (1.19%) |
| **Q10. Blood banks are facing an acute shortage these days. Would you like to be a blood donor?[Multiple responses, N=168]** |  |
| Yes | 126 (75.00%) |
| No | 42 (25.00%) |
| **Q11. Have you contributed towards the pandemic in any way yet?[Multiple responses, N=168]** |  |
| Financially | 81 (48.21%) |
| Feeding the poor | 50 (29.76%) |
| Tele consulting | 86 (51.19%) |
| Going to job regularly (essential services) | 116 (69.05%) |
| Volunteer work | 26 (15.48%) |
| Education | 71 (42.26%) |
| Other (please specify) | 6 (3.57%) |
| **Q12. How many patients have you provided tele-consulting yet?[N=13]** |  |
| 1-10 | 8 (61.54%) |
| 11-20 | 2 (15.38%) |
| 21-50 | 2 (15.38%) |
| 51-100 | 1 (7.69%) |
| >100 | 0 (0.00%) |
| **Q13. Do you think you have contributed enough?[N=91]** |  |
| Yes | 36 (39.56%) |
| No | 34 (37.36%) |
| I am not sure | 21 (23.08%) |
| **Q14. What concerns do you have regarding the current strategy the Government has employed?[Multiple responses, N=91]** |  |

**Supplementary Table 2:** Survey respondent’s suggestions on how private practitioners can contribute on a professional front.

|  |  |
| --- | --- |
| Government authorities have not involved the private sector adequately. | 53 (58.24%) |
| Private sector is involved too much, which could lead to a decrease in the healthcare given to patients of other conditions. | 6 (6.59%) |
| Government does not have enough funding. | 27 (29.67%) |
| Private sector is not part of the decision making process. | 41 (45.05%) |
| I think they are handling it perfectly | 18 (19.78%) |
| Other (please specify) | 11 (12.09%) |
| **Suggestions** | **N=41** |
| **Encourage moral values amongst the HCWs** |  |
| Honesty | 1 |
| Volunteer for COVID-19 patient care | 1 |
| **Encourage more communication between the government and private sector** |  |
| More help from the government to medical staff | 1 |
| Proper coordination | 1 |
| **Educate public** |  |
| Explain safety measures to own family and patients | 3 |
| Educate oneself about covid-19 in order to give advice to those who require it | 4 |
| Dispel myths | 2 |
| **Research** |  |
| Study patients currently under our care | 1 |
| **Train HCWs** |  |
| Non urgent patient care taught to HCWs so that experienced individuals can treat severe COVID patients | 2 |
| **Donations** | 1 |
| **Patient care** |  |
| Treat COVID patients | 9 |
| Practice Telemedicine | 7 |
| Continue practice | 5 |
| Provide outstation medical services | 1 |
| Provide treatment free of cost for the poor | 1 |
| Formulate practical protocols for patient care | 1 |

| **practitioners can contribute on a professional front.** |  |
| --- | --- |
| **Suggestions** | **N=41** |
| **Encourage moral values amongst the HCWs** |  |
| Honesty | 1 |
| Volunteer for covid patient care | 1 |
| **Encourage more communication between the government and private sector** |  |
| More help from the government to medical staff | 1 |
| Proper coordination | 1 |
| **Educate public** |  |
| Explain safety measures to own family and patients | 3 |
| Educate oneself about covid-19 in order to give advice to those who require it | 4 |
| Dispel myths | 2 |
| **Research** |  |
| Study patients currently under our care | 1 |
| **Train HCWs** |  |
| Non urgent patient care taught to HCWs so that experienced individuals can treat severe covid patients | 2 |
| **Donations** | 1 |
| **Patient care** |  |
| Treat COVID patients | 9 |
| Practice Telemedicine | 7 |
| Continue practice | 5 |
| Provide outstation medical services | 1 |
| Provide treatment free of cost for the poor | 1 |
| Formulate practical protocols for patient care | 1 |

## References

1. Nicola M, Alsafi Z, Sohrabi C, et al. The socio-economic implications of the coronavirus pandemic (COVID-19): A review. Int J Surg. 2020;78:185–193. doi:10.1016/j.ijsu.2020.04.018

2. COVID-19 to trigger roughly 195 million job losses: ILO. Accessed May 23, 2020. https://www.aa.com.tr/en/latest-on-coronavirus-outbreak/covid-19-to-trigger-roughly-195-million-job-losses-ilo/1795994

3. Guest JL, del Rio C, Sanchez T. The Three Steps Needed to End the COVID-19 Pandemic: Bold Public Health Leadership, Rapid Innovations, and Courageous Political Will. JMIR Public Health Surveill. 2020;6(2):e19043. doi:10.2196/19043

4. #IndiaFightsCorona COVID-19. MyGov.in. Published March 16, 2020. Accessed May 28, 2020. https://mygov.in/covid-19/

5. Bajpai V. The Challenges Confronting Public Hospitals in India, Their Origins, and Possible Solutions. Adv Public Health. 2014;2014:1–27. doi:10.1155/2014/898502

6. Pandey P, Sharma S. In the dark even after a decade! A 10-year analysis of India’s National Rural Health Mission: Is family medicine the answer to the shortage of specialist doctor in India? J Fam Med Prim Care. 2017;6(2):204. doi:10.4103/jfmpc.jfmpc_254_16

7. India Now Ranks Fourth Globally on Daily Increase in COVID-19 Cases. The Wire. Accessed May 27, 2020. https://thewire.in/health/india-covid-19-cases-daily-increase

8. COVID-19 | Is India’s health infrastructure equipped to handle an epidemic? Accessed May 27, 2020. https://www.brookings.edu/blog/up-front/2020/03/24/is-indias-health-infrastructure-equipped-to-handle-an-epidemic/

9. Sembiah S, Paul B, Dasgupta A, Bandyopadhyay L. Capacity Building of Private Sector Workforce for Public Health Services in India: Scope and Challenges. Indian J Community Med Off Publ Indian Assoc Prev Soc Med. 2018;43(3):144–147. doi:10.4103/ijcm.IJCM_316_17

10. Sengupta A, Nundy S. The private health sector in India. BMJ. 2005;331(7526):1157–1158. doi:10.1136/bmj.331.7526.1157

11. Loh LC, Ugarte-Gil C, Darko K. Private sector contributions and their effect on physician emigration in the developing world. Bull World Health Organ. 2013;91(3):227–233. doi:10.2471/BLT.12.110791

12. Gupta I, Bhatia M. The Indian Health Care System | Health Care | Public Health. International Healthcare System Profiles. Accessed May 27, 2020. https://international.commonwealthfund.org/countries/india/

13. Iyer V, Sidney K, Mehta R, Mavalankar D, De Costa A. Characteristics of private partners in Chiranjeevi Yojana, a public-private-partnership to promote institutional births in Gujarat, India – Lessons for universal health coverage. Dangal G, ed. PLOS ONE. 2017;12(10):e0185739. doi:10.1371/journal.pone.0185739

14. Ministry of Health and Family Welfare, Government of India. National Health Policy 2017. Accessed May 20, 2020. https://www.nhp.gov.in/NHPfiles/national_health_policy_2017.pdf

15. Eyerkaufer ML, Lima FS, Gonçalves MB. Public and private partnership in disaster risk management. Jàmbá J Disaster Risk Stud. 2016;8(1):10 pages. doi:10.4102/jamba.v8i1.277

16. Auzzir ZA, Haigh RP, Amaratunga D. Public-private Partnerships (PPP) in Disaster Management in Developing Countries: A Conceptual Framework. Procedia Econ Finance. 2014;18:807–814. doi:10.1016/S2212-5671(14)01006-5

17. Ahmed S, Gupta L. Perception about social media use by rheumatology journals: survey among the attendees of IRACON 2019. Indian J Rheumatol. doi:10.4103/injr.injr_15_20

18. Eysenbach G. Improving the Quality of Web Surveys: The Checklist for Reporting Results of Internet E-Surveys (CHERRIES). J Med Internet Res. 2004;6(3):e34. doi:10.2196/jmir.6.3.e34

19. Indian Council of Medical Research, New Delhi. Accessed May 28, 2020. https://www.icmr.gov.in/

20. Coronavirus Update (Live): 5,847,021 Cases and 359,570 Deaths from COVID-19 Virus Pandemic - Worldometer. Accessed May 28, 2020. https://www.worldometers.info/coronavirus/?utm_campaign=homeAdvegas1?%20%3Ca%20href=

21. Population, total - India | Data. Accessed May 28, 2020. https://data.worldbank.org/indicator/SP.POP.TOTL?locations=IN

22. De Ceukelaire W, Bodini C. We Need Strong Public Health Care to Contain the Global Corona Pandemic. Int J Health Serv. Published online March 18, 2020:002073142091672. doi:10.1177/0020731420916725

23. India turns to private sector to boost health coverage. Devex. Published May 1, 2017. Accessed May 23, 2020. https://www.devex.com/news/sponsored/india-turns-to-private-sector-to-boost-health-coverage-90006

24. Mills A, Brugha R, Hanson K, McPake B. What can be done about the private health sector in low-income countries? Bull World Health Organ. 2002;80(4):325–330.

25. Asia-Pacific, India. How Technology Is Changing Health Care in India. Knowledge@Wharton. Accessed May 28, 2020. https://knowledge.wharton.upenn.edu/article/technology-changing-health-care-india/

26. Armocida B, Formenti B, Ussai S, Palestra F, Missoni E. The Italian health system and the COVID-19 challenge. Lancet Public Health. 2020;5(5):e253. doi:10.1016/S2468-2667(20)30074-8

27. Spain nationalises all private hospitals, UK rents hospital beds. Accessed May 23, 2020. https://publicservices.international/resources/news/spain-nationalises-all-private-hospitals-uk-rents-hospital-beds?id=10645&lang=en.

28. Serper M, Volk ML. Current and Future Applications of Telemedicine to Optimize the Delivery of Care in Chronic Liver Disease. Clin Gastroenterol Hepatol. 2018;16(2):157- 161.e8. doi:10.1016/j.cgh.2017.10.004

29. Census of India: Literacy And Level of Education. Accessed May 28, 2020. https://censusindia.gov.in/Census_And_You/literacy_and_level_of_education.aspx

30. Bhan N, Madhira P, Muralidharan A, et al. Health needs, access to healthcare, and perceptions of ageing in an urbanizing community in India: a qualitative study. BMC Geriatr. 2017;17(1):156. doi:10.1186/s12877-017-0544-y

31. Nandan SS& R. Local ventilators need of the hour to prepare for COVID-19. Livemint. Published March 27, 2020. Accessed May 28, 2020. https://www.livemint.com/news/india/local-ventilators-need-of-the-hour-to-prepare-for-covid-19-11585332113169.html

32. Lockdown 3.0: Over 800 migrant workers reach Lucknow from Nashik by special train - The Economic Times Video | ET Now. Accessed May 28, 2020. https://economictimes.indiatimes.com/news/politics-and-nation/lockdown-3-0-over-800-migrant-workers-reach-lucknow-from-nashik-by-special-train/videoshow/75514519.cms

33. Bhutta ZA, Basnyat B, Saha S, Laxminarayan R. Covid-19 risks and response in South Asia. BMJ. 2020;368:m1190. doi:10.1136/bmj.m1190

34. Kumar S, Quinn SC. Existing health inequalities in India: informing preparedness planning for an influenza pandemic. Health Policy Plan. 2012;27(6):516–526. doi:10.1093/heapol/czr075

